# Impact of self-reported interoceptive abilities on postoperative pain

**DOI:** 10.64898/2026.10.08.26365039

**Authors:** Barbara P. Weldam, Larissa L. Meijer, Oscar F.C. van den Bosch, Marjelle E.C. van Driel, Leon Timmerman, Jacqueline F.M. van Dijk, Madelijn Strick, IPOP research group, Mienke Rijsdijk

## Abstract

Interoception is described as the sensitivity to internal bodily stimuli, and plays an important role in pain perception. Heightened interoceptive sensibility (i.e. self-reported interoceptive abilities) may reinforce misinterpretation of stimuli, enhancing pain perception. This study aimed to investigate the relationship between interoceptive sensibility and acute postoperative pain. Interoceptive sensibility was measured using the Private Body Consciousness Scale in two prospective observational surgical cohorts: a Caesarean delivery cohort (C-section cohort) of 146 patients, and a lower extremity orthopaedic surgery cohort (Ortho cohort) of 70 patients. One day after surgery, patients rated pain (primary outcome) and other postoperative symptoms (secondary outcomes) on a 0-10 scale. Bonferroni corrections were applied to secondary outcomes. Interoception positively correlated with pain (C-section cohort: Spearman’s rho = 0.182, *P* = 0.028; Ortho cohort: rho = 0.259, *P* = 0.031), and with dizziness (C-section cohort: rho = 0.309, *P* = 0.002; Ortho cohort: rho = 0.348 *P* = 0.041) in both cohorts. In the C-section cohort, interoception correlated with shivering (rho = 0.321, *P* < 0.001). In the Ortho cohort, interoception correlated with nausea (rho = 0.366, *P* = 0.023) and helplessness (rho = 0.363, *P* = 0.026); whereas no correlation with pain catastrophising was found (rho = 0.240, *P* = 0.539). In pooled multiple linear regression analysis (*n* = 216), interoception predicted pain after controlling for age, sex and anaesthetic technique (B = 0.119, 95% CI [0.027-0.210], P = 0.011). These findings highlight the potential role of interoception in identifying patients at risk for acute postsurgical pain.

**Perspective:** This study identified associations between self-reported interoceptive abilities and acute postsurgical pain in two independent cohorts. These findings could contribute to the understanding of individual differences in pain perception, and provide an initial foundation for exploring the effectiveness of interoceptive therapies in preventing severe postoperative pain.

## Introduction

Effective management of acute postsurgical pain is of great importance, as inadequate pain control negatively impacts patient outcomes (Small & Laycock, 2020). For example, it may lead to a stress response with a subsequent risk of cardiac or gastrointestinal complications (Wells et al., 2008). Furthermore, the intensity of acute postsurgical pain is an important predictor of chronic postsurgical pain (Rosenberger & Pogatzki-Zahn, 2022). The management of acute postoperative pain remains suboptimal in a significant proportion of patients (Gan, 2017). A way to improve postoperative pain management could be to gain a better understanding of why pain intensity varies between patients. The relationship between pain and interoception may provide valuable insights.

Interoception is described as the interpretation, integration and regulation of bodily stimuli by the nervous system (Crucianelli & Ehrsson, 2022). This regards internal stimuli such as heartbeat and hunger, as well as signals deriving from the body’s surface, including both noxious and pleasant stimuli (Crucianelli & Ehrsson, 2022). Interoception can be divided into three dimensions: interoceptive accuracy, interoceptive sensibility and interoceptive awareness (Garfinkel et al., 2014). Interoceptive accuracy refers to the objective ability to accurately detect signals from within the body. Interoceptive sensibility is defined as the self-perceived tendency and ability to focus on bodily states. Lastly, interoceptive awareness refers to metacognitive awareness of interoceptive accuracy. Together these dimensions represent an individual’s ability to adequately interpret bodily signals.

As interoception is linked to the integration of noxious stimuli, there is growing interest in understanding the relationship between interoception and pain (Horsburgh et al., 2024). In order to understand the relationship between pain and interoception it is necessary to look into the neurophysiological mechanisms underlying these constructs (Strigo & Simmons, 2025). Interoceptive stimuli originate from various afferent pathways, including small-diameter C and Aδ fibres, which project to lamina I of the spinal cord (Craig, 2002). Through spinothalamic pathways, several cortical areas are activated of which the insular cortex and anterior cingulate cortex (ACC) are directly linked to interoception (Berntson & Khalsa, 2021; Craig, 2002). Overlapping brain areas are involved in both pain processing and interoception (Figure 1), suggesting shared (patho)physiological pathways.

**Figure 1.**
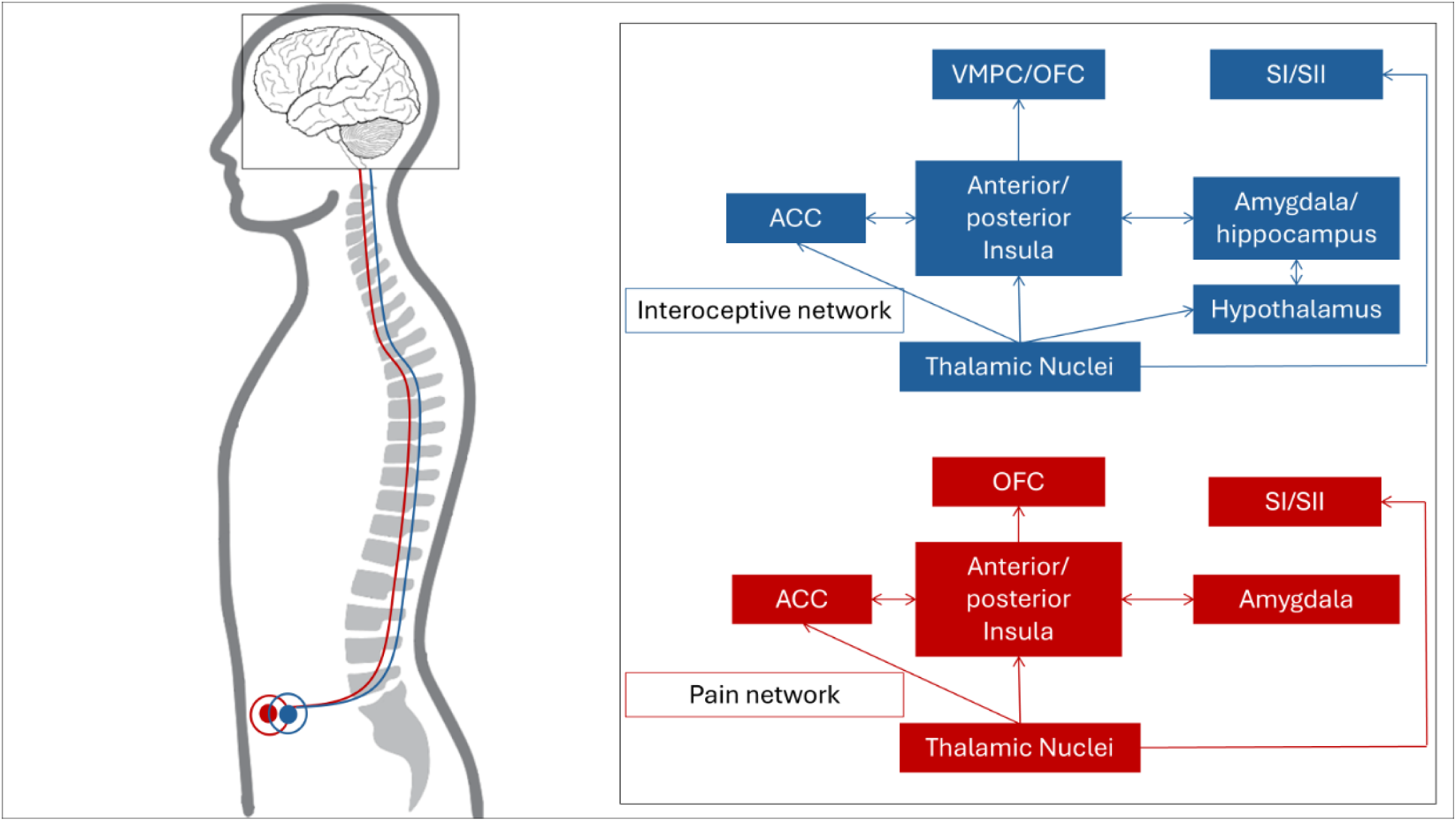
Schematic overview of neuronal projections involved in interoception and pain processing. ACC = anterior cingulate cortex; OFC = orbitofrontal cortex; VMPC = ventral medial frontal cortex; SI = primary somatosensory cortex; SII= secondary somatosensory cortex. This figure is based on the following literature: Berntson & Khalsa, 2021; Meijer et al., 2022.

Interoception is also defined as the body-brain axis of sensations, with the insula and ACC integrating interoceptive stimuli with affective and emotional states (Critchley & Patchitt, 2024; Crucianelli & Ehrsson, 2022; Strigo & Simmons, 2025). It appears that this affective component is strongly related to the interpretation of sensory input. For instance, a negative affective state (i.e. feeling sad) enhances activity in the insula and ACC, thereby priming the brain for enhanced pain perception. Furthermore, enhanced pain perception can result in a negative affective state. This feedback loop shows the important role of interoception in pain perception (Critchley & Patchitt, 2024; Strigo & Simmons, 2025).

There are individual differences in interoception, meaning that some individuals are more sensitive or aware of physiological changes than others (Duquette, 2017). Individuals with heightened interoceptive sensibility, appear to be more vulnerable to misinterpreting signals, which can intensify the pain experience (Horsburgh et al., 2024). Prior studies have focused on the relationship between chronic pain and interoception, with increased interoceptive sensibility being frequently reported among patients with chronic pain (Horsburgh et al., 2024). To date, only one study involving dancers assessed the relationship between interoception and acute pain and found no association (Delafin et al., 2022). Due to the highly specific study population of dancers, generalisability is limited. As it remains poorly understood why patients’ postoperative pain experience varies greatly and interoception appears to be related to pain perception, it is necessary to further investigate if interoceptive sensibility contributes to enhanced acute postoperative pain intensity.

By measuring interoceptive sensibility in two distinct surgical cohorts, women undergoing Caesarean delivery and patients undergoing lower extremity orthopaedic surgery, this study aims to assess if there is a relationship between interoceptive sensibility and acute postoperative pain. We hypothesize that heightened interoceptive sensibility will negatively impact postoperative pain intensity. Several treatment strategies exist that target interoception, including mindfulness-based therapies, behavioural therapies (Interoceptive Exposure, Acceptance and Commitment Therapy), and body-oriented therapies (touch interventions) (Gnall et al., 2024; Heim et al., 2023; Parma et al., 2024). Understanding these interactions could therefore be of value in the improvement of postoperative pain management.

## Methods

Data were collected during two distinct prospective cohort studies: a Caesarean delivery cohort study (C-section cohort), and an orthopaedic surgery cohort study (Ortho cohort). Both protocols have been previously published (van den Bosch et al., 2025; van Driel et al., 2025). Both studies were conducted in accordance with the Declaration of Helsinki, GCP guidelines and have ethical approval by a Medical Research Ethics Committee (Ortho cohort – NedMEC protocol number NL77085.041.21) or underwent an independent quality check to ensure compliance with legislation and regulations regarding data management, informed consent and privacy aspects (C-section cohort – protocol number 23U-0149). Only adult patients capable of understanding and providing written informed consent were included. All subjects were given the opportunity to object to the use of routinely collected data for research purposes. Further cohort-specific details are provided below. The present study is reported in accordance with the STROBE guidelines (Vandenbroucke et al., 2007). Patients or the public were not involved in the design, conduct, or dissemination of this study.

### Caesarean delivery cohort (C-section cohort)

This observational single-centre before-after study including patients undergoing Caesarean delivery under spinal anaesthesia between December 2022 until October 2023 was conducted at the Wilhelmina Children’s Hospital part of the University Medical Centre Utrecht (UMC Utrecht), The Netherlands. The study was designed to assess the impact of implementing intrathecal morphine for Caesarean delivery on postoperative quality of recovery. Patients who underwent Caesarean delivery under spinal anaesthesia were routinely asked to fill out the Obstetric Quality of Recovery questionnaire (ObsQoR-10) and Private Body Consciousness Scale (PBCS) questionnaire (Miller et al., 1981) 24 [18 to 36] hours after Caesarean delivery. The main study results have been published elsewhere (van den Bosch et al., 2025).

### Orthopaedic surgery cohort (Ortho cohort)

The second cohort originates from an ongoing prospective observational study at the UMC Utrecht and St. Antonius Hospital in Utrecht, the Netherlands, including adult patients undergoing elective lower extremity orthopaedic surgery. The study was designed to identify sensory alterations and changes in the immunological response that are related to the development of chronic postsurgical pain with neuropathic features. For the purpose of the current study, patients enrolled between September 2021 and August 2025 were included. Preoperative questionnaires were completed, including the PBCS. This questionnaire was added through a protocol amendment after initiation of the study. One day after surgery, the International Pain Outcome Questionnaire (IPOQ) was completed (Rothaug et al., 2013). A detailed description of methodology can be found in the published protocol (van Driel et al., 2025).

### Measurements and data handling

In both cohorts, Interoceptive sensibility was assessed using the <u>Private Body Consciousness</u> <u>Scale (PBCS)</u>, a subscale of The Body Consciousness Questionnaire (Miller et al., 1981). In this 5-item questionnaire, patients rate awareness of internal sensations on 5-point Likert scales (1 = extremely uncharacteristic – 5 = extremely characteristic), yielding a total score of 5-25. Standardization for interoceptive sensibility is 12.0 (SD 3.3) for women and 11.7 (SD 3.0) for men (Miller et al., 1981).

In the C-section cohort, patients completed the <u>Obstetric Quality of Recovery</u> <u>(ObsQor-10) questionnaire</u> 24 [18 to 36] hours after surgery (van den Bosch et al., 2025). In this 10-item questionnaire, patients rate several items on postpartum recovery (e.g. pain, nausea and vomiting, dizziness, and shivering) on an 11-point numeral rating scale (NRS). The first item reported average pain over the past 24 hours, and was formulated as follows: “Please score the severity of the below symptoms (*pain*) in the past 24 hours”. Furthermore, alongside the ObsQor-10, patients rated their general health on a visual analogue scale (GH-VAS; 0 = worst – 100 = best health state).

In the Ortho cohort, patients completed the <u>International Pain Outcome</u> <u>Questionnaire (IPOQ)</u> one day after surgery (Rothaug et al., 2013). This questionnaire consists of 13 main items that assess several aspects of pain-related recovery, including side effects (nausea, drowsiness, itching, dizziness) and emotional interference (anxiety, helplessness). An item measuring the pain score on an 11-point NRS at the time of completion was added to the IPOQ, which was formulated as follows: “On this scale, please indicate the severity of pain you’re experiencing at this moment”.

Furthermore, in the Ortho cohort, patients completed the <u>Pain Catastrophising Scale (PCS)</u> at baseline (Van Damme et al., 2002). This 13-item questionnaire uses 5-point Likert scales (0 = not at all – 4 = all the time) to measure negative pain-related thoughts and feelings, yielding a total score of 0-52. The questionnaire comprises three dimensions of pain catastrophising: rumination, magnification and helplessness.

<u>Patient characteristics</u> were extracted from the Electronic Health Record, including patient age, sex, BMI, history of chronic pain (self-reported in the Ortho cohort), opioid use and ASA classification. Data on anaesthetic technique, and analgesic regimen were extracted from the perioperative records.

### Endpoints

The primary endpoint of this study was acute postoperative pain, defined as the patient-reported NRS 24 hours after surgery. Secondary endpoints included bodily stimuli measured by the ObsQor-10 or IPOQ, a priori selected as relevant to interoception (nausea, vomiting, dizziness, shivering, drowsiness, itching). Further secondary endpoints were: total ObsQor-10 score, GH-VAS, PCS, and the emotional interference of pain (anxiety and helplessness scores on the IPOQ).

### Statistical analysis

No statistical power calculation was conducted prior to the study. The sample size was determined by the data available across both cohorts. Missing data were addressed by restricting analyses to cases with complete data for the parameter under investigation. The number of missing values was reported accordingly. Normality was assessed using the Shapiro-Wilk test and visual inspection of histograms and Q-Q plots. Continuous normally distributed data are expressed as mean (SD), otherwise, they are presented as median (IQR). Categorical data are presented as frequencies (%).

Exploratory analyses of the relationship between interoception and pain included Spearman’s correlation coefficients accompanied by scatterplots with LOESS curves for visual evaluation, and subgroup analyses stratified by sex (Grabauskaitė et al., 2016). For the secondary outcome measures, Spearman’s correlations were calculated, with *P* values adjusted using Bonferroni correction. Spearman’s correlation coefficients of 0.1-0.3 were considered weak, 0.4-0.6 moderate, and 0.7-0.9 strong. Additionally, *post hoc* third-order regression with orthogonal polynomials was conducted on pooled data, to explore a non-linear pattern.

To evaluate the hypothesized relationship, simple linear regression analyses were performed per study, followed by multiple linear regression analyses on the data of all studies combined to adjust for potential confounders. Confounding variables included were age and sex (Grabauskaitė et al., 2016; Pfeifer & Cawkwell, 2024; Thurston et al., 2022; van Dijk et al., 2021), with anaesthetic technique and surgery type as covariates (El-Tallawy et al., 2025; Papadomanolakis-Pakis et al., 2025). Multicollinearity was assessed using variation inflation factors (VIF); variables with VIF > 5 were excluded from the model. Results are presented as unstandardised regression coefficients (*B*) with confidence intervals (CI) and *P* values, alongside adjusted *R*^2^ and standardized regression coefficients (β) of the multiple regression analysis.

Significance was set at *P ≤* 0.05. Statistical analyses were performed using IBM SPSS Statistics version 30.0 and R version 4.5.1.

## Results

A total of 216 patients were included across two cohorts. In the C-section cohort, a total of 158 patients were approached, of who 146 completed both questionnaires. In the Ortho cohort, of 578 eligible patients, 104 were included, of whom 70 provided complete responses (n = 28 missing due to protocol amendment).

### Patients’ characteristics

Baseline characteristics for each study are presented in Table 1.

**Table 1.**
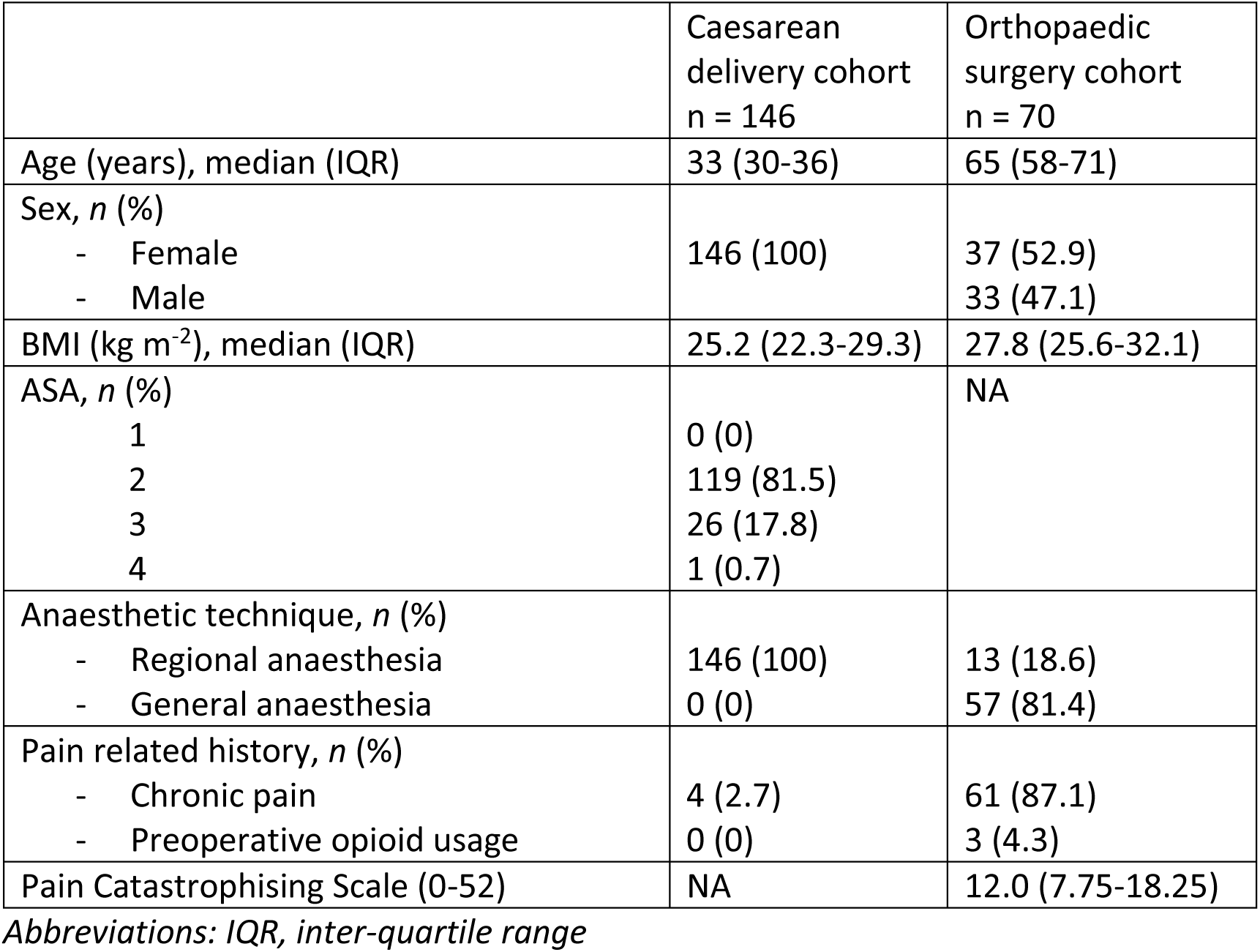
Baseline patient, surgical and anaesthetic characteristics.

### Correlation of interoceptive sensibility and postoperative pain

In the C-section cohort, the postoperative mean PBCS score was 15.5 (SD 3.6) in females. In the Ortho cohort, the preoperative mean PBCS scores were: 13.5 (SD 4.5) in females, 12.4 (SD 2.9) in males, and 13.0 (SD 3.9) in total (Supplementary Table S1).

The postoperative median pain score after 24 hours was 5.0 (IQR 3.0-7.0) in the C-section cohort, intrathecal morphine was administered to 59 (40.4%) patients. In the Ortho cohort, the median pain score after 24 hours was 4.0 (IQR 2.0-6.25), postoperative use of opioids (morphine or oxycodone) was reported in 48 (68.6%) patients (missing data 11.4%). Further details regarding postoperative symptom scores are presented in Table 2.

**Table 2.** Perioperative symptoms and their correlation with interoceptive sensibility.

|  | Symptom scores,<br>median (IQR) | Spearman's rho<br>correlation | P value |
| --- | --- | --- | --- |
| **Caesarean delivery cohort**<br>(n = 146)<sup>a</sup> |  |  |  |
| <u>Postoperative</u> |  |  |  |
| ObsQor-10 (0-100) | 69 (58-82) | -0.142 | 1.000 |
| 1. Pain last 24h (0-10) | 5 (3-7) | 0.182 | 0.028* |
| 2. Nausea and vomiting<br>(0-10) | 0.0 (0.0-3.25) | 0.235 | 0.051 |
| 3. Dizziness (0-10) | 2 (0-5) | 0.309 | 0.002** |
| 4. Shivering (0-10) | 1 (0-4) | 0.321 | <0.001*** |
| GH-VAS (0-100) | 61 (50-75) | -0.061 | 1.000 |
| **Orthopaedic surgery**<br>**cohort**<br>(n = 70)<sup>b</sup> |  |  |  |
| <u>Preoperative</u> |  |  |  |
| Pain catastrophising scale<br>(0-52) | 12.0 (7.75-18.25) | 0.240 | 0.539 |
| <u>Postoperative</u><br>IPOQ |  |  |  |
| Pain at 24h (0-10) | 4.0 (2.0-6.25) | 0.259 | 0.031* |
| *Emotional interference* |  |  |  |
| Anxiety (0-10) | 0 (0-1) | 0.185 | 1.000 |
| Helplessness (0-10) | 2 (0-4) | 0.363 | 0.026* |
| *Side effects* |  |  |  |
| Nausea (0-10) | 0 (0-2) | 0.366 | 0.023* |
| Drowsiness (0-10) | 3 (1.0-6.5) | 0.278 | 0.248 |
| Itching (0-10) | 0 (0-0) | 0.190 | 1.000 |
| Dizziness (0-10) | 1 (0-4) | 0.348 | 0.041* |

*Note: Interoceptive sensibility was measured with the Private Body Consciousness Scale.* *Asymptotic t approximation was used to calculate P values. With the exception of pain (the* *primary outcome), P values were corrected for multiple testing (Bonferroni).*
*Abbreviations: ObsQor-10, Obstetric Quality of Recovery score; GH-VAS, general health* *measured by visual analogue score; IPOQ, International Pain Outcome Questionnaire.*
*\* P < 0.05. \*\* P < 0.01. \*\*\* P < 0.001.*
*<sup>a</sup> ObsQor-10 and GH-VAS n = 139.*
*<sup>b</sup> Pain Catastrophising Scale and pain n = 70, other symptoms n=69.*

A weak yet significant positive correlation between the PBCS and acute postoperative pain was identified using Spearman’s rho in both cohorts (Table 2; Fig. 2). As for the secondary outcome measures, in the C-section cohort, weak yet significant positive correlations with the PBCS were observed for dizziness and shivering. The total score of the ObsQor-10 and the GH-VAS did not correlate with the PBCS. In the Ortho cohort, nausea, dizziness and helplessness correlated with the PBCS, whereas the PCS, anxiety, drowsiness and itching did not.

**Figure 2.**
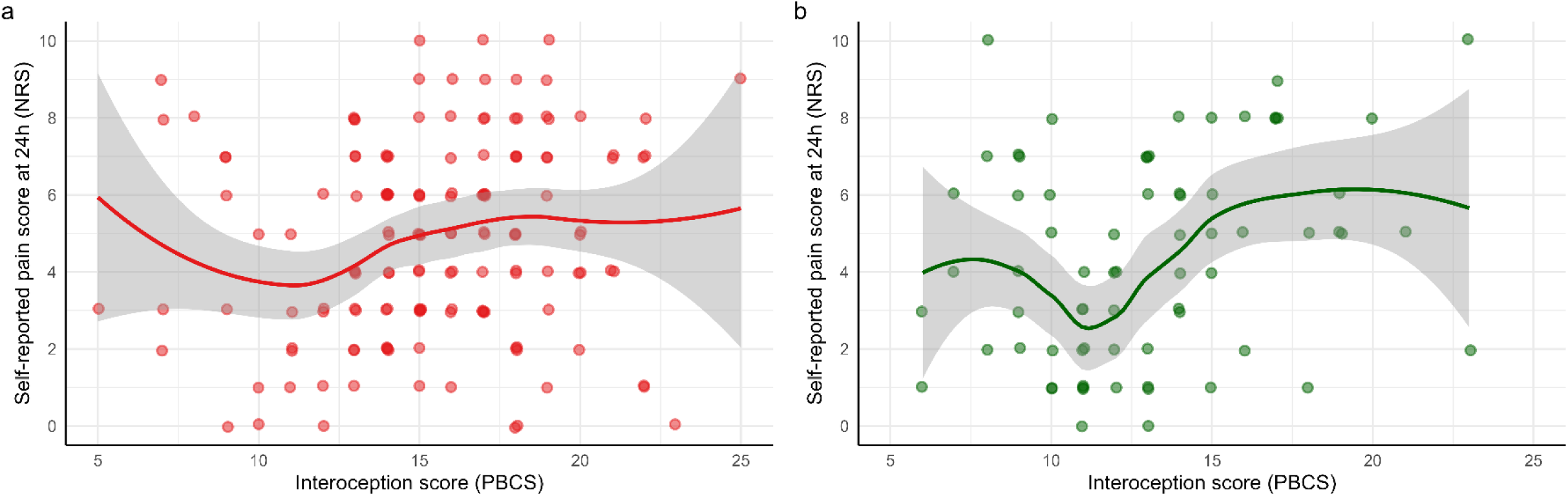
Scatterplots with LOESS curves illustrating the relationship between interoceptive sensibility and acute postsurgical pain. Interoceptive sensibility is shown on the x-axis, assessed with Private Body Consciousness Scale (PBCS). Acute postoperative pain (NRS) is shown on the y-axis. Panel a depicts an all-female cohort (caesarean delivery, n = 146). Panel b (orthopaedic surgery, n = 70) shows a mixed-sex cohort. Pain was measured 24 hours after surgery. A LOESS regression line (span = 0.75) is shown in red (caesarean delivery cohort), and green (orthopaedic surgery cohort), with 95% confidence intervals in grey.

In subgroup analysis of the Ortho cohort (Fig. 3), a significant moderate positive correlation (Spearman’s ρ = 0.413, *P* = 0.011) between the PBCS and pain was observed in female patients, while no significant correlation was observed in male patients (Spearman’s ρ = −0.002, *P* = 0.992).

**Figure 3.**
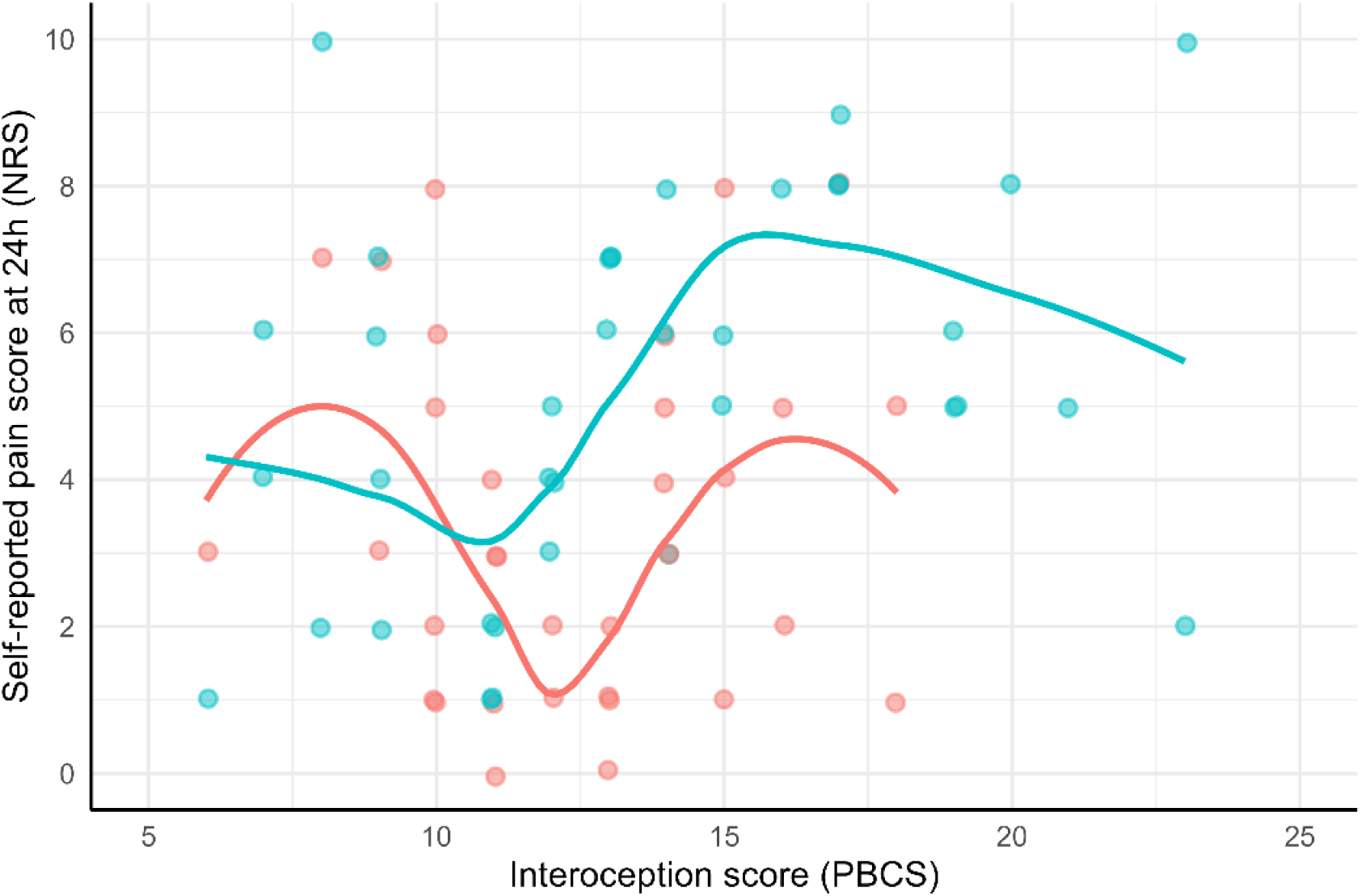
Scatterplot with LOESS curves illustrating the relationship between interoceptive sensibility and pain, stratified by sex. Subgroup analysis of the lower extremity orthopaedic surgery cohort, with LOESS regression lines (span = 0.75) in blue (females, n = 37) and orange (males, n = 33). Interoceptive sensibility is shown on the x-axis, assessed with Private Body Consciousness Scale (PBCS). Acute postoperative pain (NRS), measured 24 hours after surgery, is shown on the y-axis.

### Predictive value of interoception for acute postoperative pain

In univariate regression analysis of the C-section cohort, interoception was not a significant predictor of pain (*B* = 0.108, 95% CI [-0.005-0.222], *P* = 0.061). In the Ortho cohort, interoception was a significant positive predictor of pain (*B* = 0.185, 95% CI [0.025-0.345], *P* = 0.024).

Combining the data of both studies (*n* = 216), interoception was a significant positive predictor of pain in both the univariate model (*B* = 0.139, 95% CI [0.052-0.227], *P* = 0.002) and after adjusting for confounders (*B* = 0.119, 95% CI [0.027-0.210], *P* = 0.011), with the multiple regression model explaining 9.2% of variance, *R*^2^ = 0.092, *F*(4,211) = 5.344, *P* < 0.001 (Table 3). Due to multicollinearity with age and anaesthetic technique, surgery type was excluded from the model (VIF = 6.841). In *post hoc* polynomial regression, the linear term was significant (*P* = 0.009), whereas the quadratic (*P* = 0.735) and cubic (*P* = 0.093) terms were not (Supplementary Figure S1).

**Table 3.**
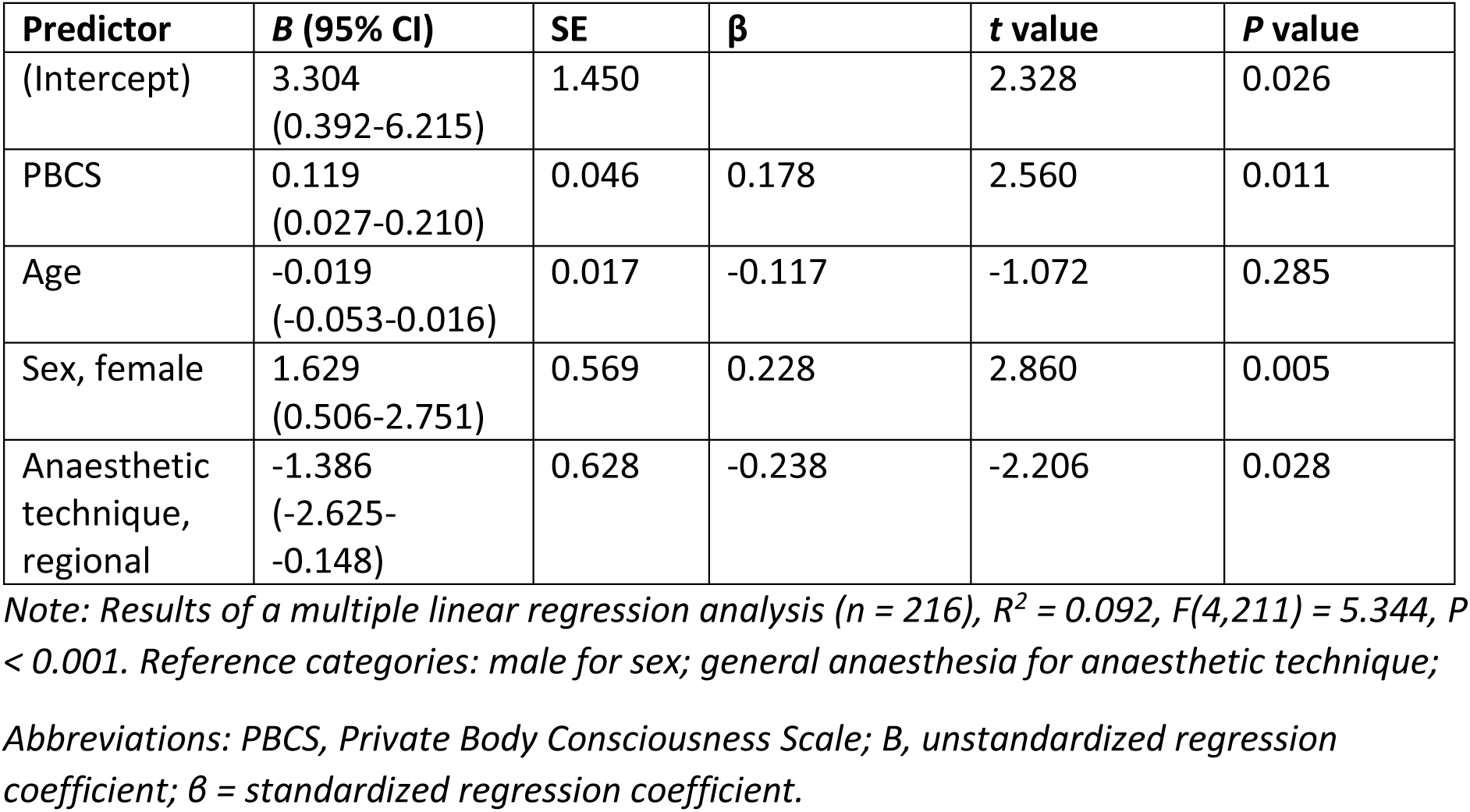
Predictive value of interoception for acute postoperative pain after adjusting for confounders and covariates.

## Discussion

This study examined the association between interoceptive sensibility and acute pain in a broad surgical population including women undergoing Caesarean delivery and patients receiving lower extremity orthopaedic surgery. A positive correlation between interoceptive sensibility and acute pain was identified in both cohorts. Combining the data of both cohorts, interoceptive sensibility predicted acute postsurgical pain after controlling for age, sex and anaesthetic technique. Positive correlations were additionally identified between interoception and dizziness, shivering, nausea and helplessness; no correlations were observed with pain catastrophising and anxiety. Subgroup analysis of the Ortho cohort revealed that interoception was associated with acute pain in female patients, but not in male patients. The exploratory analyses suggested a non-linear association between interoceptive sensibility and acute pain; further analysis did not support this pattern, suggesting that the association is most likely linear.

The findings of this study suggest that heightened interoceptive sensibility may play a role in enhanced pain perception. A possible explanation is that this heightened interoceptive sensibility is accompanied by lowered interoceptive accuracy, meaning that while patients are more attentive to bodily signals, they are unable to accurately interpret them. This discrepancy may reflect a ‘prediction error’. It is theorised that the brain does not solely passively sense, but rather generates top-down predictions about the body’s internal state based on contextual information such as affective states or prior experience (Quadt et al., 2018; Strigo & Simmons, 2025). A prediction error occurs when there is a mismatch between sensory information and these predictions. The discrepancy between interoceptive accuracy and interoceptive sensibility may thus reflect such a prediction error, as the self-perceived interoceptive abilities (the top-down prediction) exceed the objective capacity to adequately process bodily signals (Quadt et al., 2018). Consequently, it is possible that noisy interoceptive signals may be amplified or misattributed as threatening, thereby contributing to enhanced pain perception. As the current study only measured interoceptive sensibility, this hypothesis remains speculative and warrants further investigation.

Currently, there is only one other study into acute pain and interoception, which did not find a difference in self-reported interoceptive abilities between dancers with acute pain and their pain-free controls (Delafin et al., 2022). The study included 26 dancers with an unspecified type of acute pain, with interoception assessed using the Multidimensional Assessment of Interoceptive Awareness (MAIA) questionnaire. The discrepancy in findings between the prior study and the present one, may be attributed to variations in sample size and the use of a different questionnaire, as the MAIA is more extensive than the PBCS. Interestingly, evidence suggests that trained individuals exhibit heightened interoceptive accuracy (Kleber et al., 2025; Zeng et al., 2025). Therefore, another possible explanation for the absence of an association in the study by Delafin et al. (2022) may be that dancers exhibit a smaller discrepancy between interoceptive sensibility and interoceptive accuracy.

More studies have focused on the relationship between chronic pain and interoception, which indicate that patients with chronic pain have an impaired interoceptive accuracy and enhanced interoceptive sensibility (Horsburgh et al., 2024). Our finding that higher interoceptive sensibility may be associated with higher acute pain scores could reflect the enhanced interoceptive sensibility found in patients with chronic pain. Additionally, lower scores in specific domains of the MAIA questionnaire, namely Not-Worrying, Self-Regulation and Trusting, have also been associated with chronic pain (Horsburgh et al., 2024). This suggests that not merely the degree of attention is of clinical relevance, but also the differentiation between anxiety-driven and mindful attention styles (Mehling, 2016). The current study utilised the PBCS, which does not differentiate between attention styles. Given the observed correlations with helplessness but not with anxiety and pain-catastrophising, it remains uncertain whether the PBCS reflects an anxiety-driven attention style.

There are also studies that do not show a relationship between interoception and chronic pain (d’Alcalà et al., 2014; Valenzuela-Moguillansky et al., 2017). Thus, it remains unclear whether chronic pain develops due to a disruption in interoception or dysfunctional interoception develops as a result of lingering pain (Horsburgh et al., 2024). Despite the uncertain direction of the relationship, interoception appears to play an important role in pain perception, as emphasised by the presence of shared neuroanatomical pathways (Craig, 2002; Strigo & Simmons, 2025). Interestingly, the current study shows that interoceptive sensibility can be a predictor for acute postsurgical pain. As the intensity of acute postsurgical pain is an important predictor of chronic postsurgical pain, the development of such pain may be prevented by early targeting of interoceptive abilities, preferably before surgery (van Driel et al., 2022).

Several treatment options exist that target the interoceptive system, including touch-interventions (Di Lerna et al., 2020). These can easily be provided before surgery, are relatively time-efficient and may not require a trained specialist or therapist. This contrasts with common treatment options for pain-related anxiety and pain catastrophising, such as Cognitive Behavioural Therapy or multimodal programmes (Huang et al., 2011; Michaelides & Zis, 2019; Yokoyama et al., 2024). These therapies appear quite effective, yet require several sessions and are therefore time-consuming. Furthermore, restoring interoceptive abilities is not yet included in these therapies. Future studies should therefore focus on studying the effectiveness of interoceptive treatments in reducing postsurgical pain.

Our study has strengths and limitations. A notable strength is addressing the gap in the literature concerning the association between interoception and acute pain. Another strength is the inclusion of two distinct study populations, which increases generalizability. A limitation is the usage of the PBCS, as this questionnaire provides quite a crude score for interoception. Interoceptive sensibility may be regarded as a multidimensional construct, which is not fully captured by the PBCS alone (Todd et al., 2022). Furthermore, as the two cohorts differed in study design, not all potential confounding variables and covariates could be controlled for. As a result, it is possible that the observed relationship might be partially explained by unmeasured variables. Future studies should aim to gain more knowledge on the relationship between interoception and postsurgical pain, taking the multidimensionality of interoception into account.

In conclusion, this is the first study to show a positive relationship between interoception and acute postsurgical pain, meaning that patients with higher interoceptive sensibility report higher pain scores. This was found in both surgical populations. Additionally, interoception is a predictor for postsurgical pain after controlling for age, sex, and anaesthetic technique. Current results provide an initial foundation to explore the effectiveness of interoceptive therapies in the perioperative setting, with an ultimate goal to improve pain management.

## Authors’ contributions

This study was conceptualised by MS, MR and the IPOP research group. Methodology was developed by MR, LM, MS, and BW. Data were collected by LT and JD, and curated by BW, OB, and MD. BW performed formal analysis and data visualisation, supervised by MR. The original draft of the manuscript was written by BW, LM and MR. All authors contributed to reviewing and editing the manuscript, and approved the final version. Supervision was provided by LM and MR.

## Data Availability

All data produced in the present study are available upon reasonable request to the authors

## Acknowledgements

We would like to acknowledge Suzanne E. Rosier and Lottie van Baal for their valuable contributions to data collection on the Caesarean delivery cohort, and Timme Schaap for providing the opportunity to collect data under his supervision on the maternity ward. Our sincere thanks go to the <u>Interdisciplinary Pain Research Platform–the Netherlands (IPOP-NL)</u> <u>group</u>–Sylvia Brugman, Yoeri van de Burgt, Tessa van Charldorp, Jannie de Grauw, Frank Meye, Mienke Rijsdijk, Madelijn Strick, Hanneke Willemen, and Laura Winkens—for fostering collaboration across disciplines and pioneering the introduction of interoception to the postsurgical patient population. Finally, we want to thank all the patients for their willingness to fill in the questionnaires.

## Data availability

The dataset of the C-section cohort is available upon reasonable request to the corresponding author. As the Ortho cohort originates from an ongoing study, data will be available upon request after publication of the main study findings.

## Legends for illustrations and tables

