## Supplementary Table S1, Supplementary Figure S1 for "Impact of self-reported interoceptive abilities on postoperative pain"

**Supplementary material**

**Supplementary table S1.** Complete overview of interoceptive sensibility scores per cohort, as measured with the Private Body Consciousness Scale.

| | **Caesarean delivery cohort** (n = 146) | **Lower extremity orthopaedic surgery cohort** (n = 70) | | |
|---|---|---|---|---|
| | Female | Male | Female | Total |
| Total body awareness score, *mean (SD)* | 15.5 (3.6) | 12.4 (2.9) | 13.5 (4.5) | 13.0 (3.9) |
| I am sensitive to internal bodily tensions, *median (IQR)* | 3 (2 to 4) | 3 (2-3) | 3 (2-4) | 3 (2-3) |
| I know immediately when my mouth or throat gets dry, *median (IQR)* | 5 (3,75 to 5) | 3 (2-4) | 2 (2-4) | 2.5 (2- 4) |
| I can often feel my heart beating, *median (IQR)* | 2 (1 to 3) | 2 (2-3) | 3 (2-3) | 2 (2-3) |
| I am quick to sense the hunger contractions of my stomach, *median (IQR)* | 3 (2 to 4) | 2 (1.5-3) | 3 (2-3.5) | 2 (2-3) |
| I'm very aware of changes in my body temperature, *median (IQR)* | 3 (2 to 4) | 2 (2-3) | 3 (1-4) | 2.5 (1.0-3.25) |

**Supplementary figure S1.** Results of cubic polynomial regression analysis

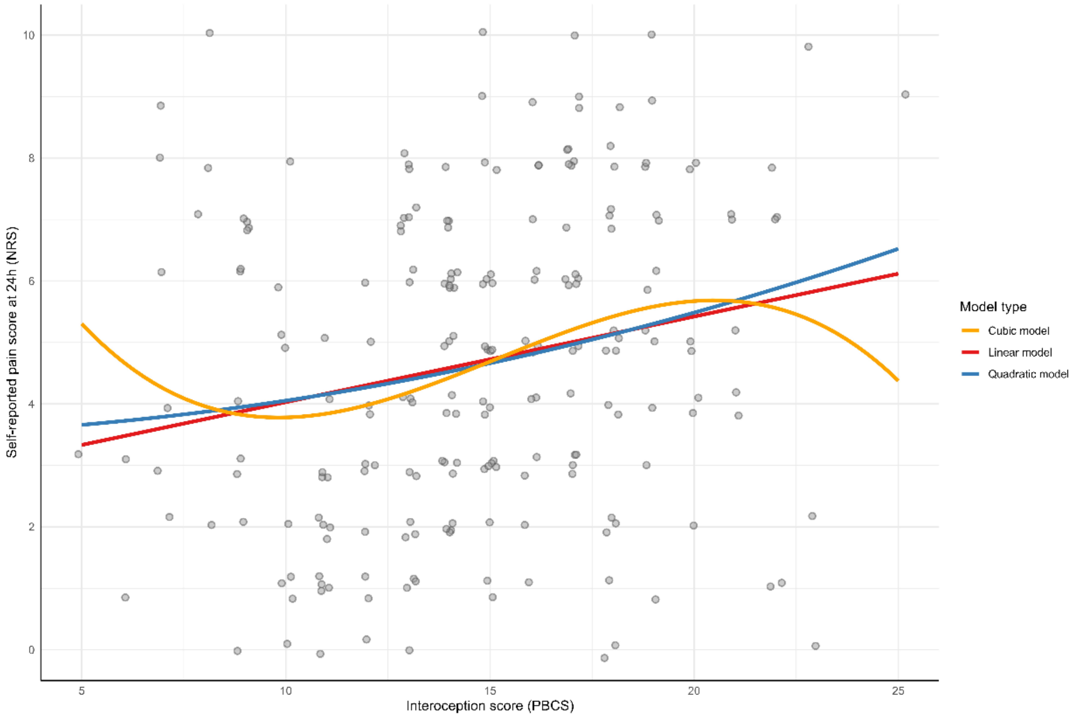


The figure depicts the linear (red; pain ~ PBCS), quadratic (blue; pain ~ PBCS + I(PBCS^2)) and cubic (orange; pain ~ PBCS + I(PBCS^2) + I (PBCS^3)) models predicting acute postoperative pain (NRS) from interceptive sensibility as measured with the Private Body Consciousness Scale (PBCS). Results of the polynomial regression were as follows:

```
Call:
lm(formula = pain ~ poly(PBCS, 3) + sex + age + anaesthetic_technique,
    data = pooled_interoception)

Residuals:
    Min      1Q  Median      3Q     Max 
-5.5204 -1.8789 -0.2441  1.7796  5.5002 

Coefficients:
                               Estimate Std. Error t value Pr(>|t|)    
(Intercept)                     5.17662    1.23848   4.180 4.29e-05 ***
poly(PBCS, 3)1                  6.95227    2.63073   2.643  0.00885 ** 
poly(PBCS, 3)2                  0.84311    2.49203   0.338  0.73546    
poly(PBCS, 3)3                 -4.22895    2.50292  -1.690  0.09259 .  
sexfemale                       1.54837    0.57178   2.708  0.00733 ** 
age                            -0.01803    0.01746  -1.033  0.30298    
anaesthetic_techniqueRegional -1.41979    0.62714  -2.264  0.02461 *  
---
Signif. codes:  0 '***' 0.001 '**' 0.01 '*' 0.05 '.' 0.1 ' ' 1

Residual standard error: 2.473 on 209 degrees of freedom
Multiple R-squared:  0.1047,	Adjusted R-squared:  0.07902 
F-statistic: 4.074 on 6 and 209 DF,  p-value: 0.0006909
```
